# Base Reproduction Number of COVID-19: Statistic Analysis

**DOI:** 10.1101/2020.09.26.20202010

**Authors:** Hongjun Zhu, Jiangping Huang, Xin Liu

## Abstract

The coronavirus disease 2019 (COVID-19) has grown up to be a pandemic within a short span of time. The quantification of COVID-19 transmissibility is desired for purposes of assessing the potential for a place to start an outbreak and the extent of transmission in the absence of control measures. It is well known that the transmissibility can be measured by reproduction number. For this reason, the large amount of research focuses on the estimations of reproduction number of COVID-19. However, these previous results are controversial and even misleading. To alleviate this problem, Liu et al advised to use averaging technique. Unfortunately, the fluctuant consequence principally arises from data error or model limitations rather than stochastic noise, where the averaging technique doesn’t work well. The most likely estimation in USA and Wuhan is about 8.21 and 7.9. However, no enough evidence demonstrates the transmissibility increase of infectious agent of COVID-19 throughout the world.

## Background

The coronavirus disease 2019 (COVID-19) has grown up to be a pandemic within a short span of time. The quantification of COVID-19 transmissibility is desired for purposes of assessing the potential for a place to start an outbreak and the extent of transmission in the absence of control measures[1]. It is well known that the transmissibility can be measured by reproduction number. For this reason, the large amount of research focuses on the estimations of reproduction number of COVID-19. However, these previous results are controversial and even misleading. To alleviate this problem, Liu et al[2] advised to use averaging technique. Unfortunately, the fluctuant consequence principally arises from data error or model limitations rather than stochastic noise, where the averaging technique doesn’t work well. This has inspired a collection of studies on reproduction number of COVID-19.

## Objective

To find out the reason for estimation change and then the most reliable estimation of base reproduction number.

## Definitions

Reproduction number can be subdivided into basic reproduction number and effective reproductive number. In this paper, our discussion will be confined to basic reproduction number since it is harder to calculate than the other[3]. Even so, the definitions of both are presented here for comparison.

1. Basic reproduction number (BRN) *R*_0_ is defined as the expected number of secondary infectious cases generated by an average infectious case in an otherwise uninfected population[1, 4].
2. Effective reproductive number (ERN), *R*_*e*_, is the number of secondary cases generated by an infectious case once an epidemic is underway[1].

BRN *R*_0_ can be expressed as *R*_0_=*kpd*, where *k* is the number of contacts each infectious individual has per unit time, *p* is the probability of transmission per contact between an infectious case and a susceptible individual, and *d* is the mean duration of infectiousness[1]. In the absence of control measures, *R*_*e*_=*rR*_0_, where *r* is the proportion of the population susceptible [1]. Here, *r* < 1.

## Data

Google Scholar and Science Citation Index were used to search for eligible studies about BRN of COVID-19. The estimations of BRN range from 0.3[5] to 8.213[6]. An extensive and resulting description is given in Table 1.

**Table 1.** The base reproductive number *R*_0_

| ID | Date | $R_0$ | 95% CI | Place | First author |
| --- | --- | --- | --- | --- | --- |
| 1 | 1/11 | 2.2 | 1.4-3.9 | Wuhan, China | Li [4] |
| 2 | 1/13 | 0.3 | 0.17-0.44 | Wuhan, China | Wu[5] |
| 3 | 1/15 | 2.6 | 2.49-2.63 | China | Zhao[7] |
| 4 | 1/18 | 2.2 | - | Wuhan, China | Riou[8] |
| 5 | 1/18 | 4.6 | 3.56-5.65(90%) | Hubei, China | Anastassopoulou[9] |
| 6 | 1/22 | 3.11 | 2.39-4.13 | Wuhan, China | Read[10] |
| 7 | 1/22 | 3.15 | - | China | Tian[11] |
| 8 | 1/23 | 2.9764 | - | Anhui, China | Tian[12] |
| 9 | 1/24 | 2.24 | 1.96-2.55 | China | Zhao[13] |
| 10 | 1/24 | 3.58 | 2.89-4.39 | China | Zhao[13] |
| 11 | 1/24 | 3.2 | 2.7-3.7 | Wuhan, China | Jung[14] |
| 12 | 1/24 | 2.1 | 2.0-2.2 | Wuhan, China | Jung[14] |
| 13 | 1/28 | 2.68 | 2.47-2.86 | Wuhan, China | Wu[15] |
| 14 | 1/30 | 4.7092 | - | Wuhan, China | Zhao[16] |
| 15 | 1/30 | 5.934 | - | Hubei, China | Zhao[16] |
| 16 | 1/30 | 1.5283 | - | China | Zhao[16] |
| 17 | 2/2 | 2.55 | 2.25-2.96 | China | Aghaali[17] |
| 18 | 2/8 | 2.79 | - | - | Liu[2] |
| 19 | 2/11 | 5.2 | 5.04-5.47 | Wuhan, China | Mizumoto[18] |
| 20 | 2/29 | 2.7999 | - | China | Dur-e-Ahmad[19] |
| 21 | 2/29 | 1.78 | 1.36-2.58 | South Korea | Aghaali[17] |
| 22 | 2/29 | 2.6 | 2.4-2.8 | Japan | Kuniya[20] |
| 23 | 2/29 | 3.15 | 2.41-3.90 | - | He [21] |
| 24 | 3/1 | 2.6 | 2.3-2.9 | Korea | Zhuang[22] |
| 25 | 3/1 | 3.2 | 2.9-3.5 | Korea | Zhuang[22] |
| 26 | 3/4 | 2.7 | 2.1-3.4 | Shahroud, Iran | Khosravi[23] |
| 27 | 3/5 | 2.6 | 2.3-2.9 | Italy | Zhuang[22] |
| 28 | 3/5 | 3.3 | 3.0-3.6 | Italy | Zhuang[22] |
| 29 | 3/6 | 2.71 | - | Wuhan, China | Wang[24] |
| 30 | 3/7 | 1.82 | 1.64-2.05 | Iran | Aghaali[17] |
| 31 | 3/7 | 3.47 | 3.16-3.84 | Qom, Iran | Aghaali[17] |
| 32 | 3/8 | 2.76-3.25 | - | Italy | Remuzzi[25] |
| 33 | 3/9 | 0.945 | - | Wuhan, China | Ndairoua[26] |
| 34 | 3/9 | 3.37 | 3.03-3.81 | Italy | Aghaali[17] |
| 35 | 3/10 | 7.9 | - | Wuhan, China | Zhu[27] |
| 36 | 3/13 | 3.2-3.6 | - | Wuhan, China | Davies[28] |
| 37 | 3/19 | 2.3 | - | Ontario, Canada | Tuite[29] |
| 38 | 3/23 | 1.593 | 1.582-1.604 | Korea | Xu[6] |
| 39 | 3/23 | 8.213 | 8.139-8.288 | USA | Xu[6] |
| 40 | 3/25 | 5.25 | - | Brazil | Crokidakis[30] |
| 41 | 4/3 | 2.58 | - | Costa Rica | Chaves[31] |
| 42 | 4/5 | 2.38 | - | Italy | Giordano[32] |
| 43 | 4/19 | 5.0692 | - | Heilongjiang, China | Sun[33] |
| 44 | 5/2 | 4.86 | - | Iran | Sahafizadeh[34] |

Note that CI indicates confidence interval. Date means the time when data was collected.

## Discussion

### BRN increases with time

In order to prove the BRN varies with time, the data is divided into two groups: (1) Group I consists of the first 22 estimations of BRN in Table 1; (2) Group II is composed of the last 22 estimations. A statistical hypothesis testing is carried out here. The null hypothesis is that the mean of Group I is no less than Group II. The alternative hypothesis is that the mean of Group II is more than Group I.

Suppose the estimations follows a normal distribution, if the null hypothesis is really true, the test statistic *t* which follows by the Student’s t-distribution should be more than -1.732 with probability 95%, while it is -2.256 according to the data. Therefore, the null hypothesis is rejected and the conclusion is that BRN increases with time.

### Difference between methods

For more than two categories, some of which have small numbers, statistic analysis tends to pool some of the categories together[35]. For this reason, the estimation methods of BRN are divided into three broad categories: (1) methods based on exponent-growth-like models, (2) methods based on SIR epidemic models, and (3) others. For convenience, we will hereafter use ‘EX method’ to denote the method based on exponent-growth-like models and ‘SIR method’ to indicate method based on susceptible–infectious–removed (SIR) model.

At first, we suspect there is a significant difference between the groups in variance. This is confirmed by using Levene’s test (on the means) since the Levene test statistic 12.28 is far more than 3.23, which is the upper critical value of the F distribution with 2 and 40 degrees of freedom at a significance level of 0.05.

For the reason that the variances and sample sizes are unequal across groups, Welch’s Test is used to perform an ANOVA analysis. Here, the null hypothesis is that the means of the three groups are equal. The alternative hypothesis is that more than one group is different from others. From the data, the Welch’s test statistic is 3.25, which more than the upper critical value F(2, 14.165) at a significance level of 0.1. As a result, the conclusion can be drawn that more than one group is different from others.

Further, the results of three methods are shown by points in Figure 1. Here, the estimation (3.2-3.6) provided by Davies [28] is substituted by 3.4 for the convenience of pictorial display, and 2.76-3.25 by 3.005 for the same reason. Regression analysis is implemented separately for each method, as the lines shown in Figure 1. They all show that BRN increases with time. Note that BRN 0.3 is rejected since an epidemic can occur if and only if.

**Figure 1.**
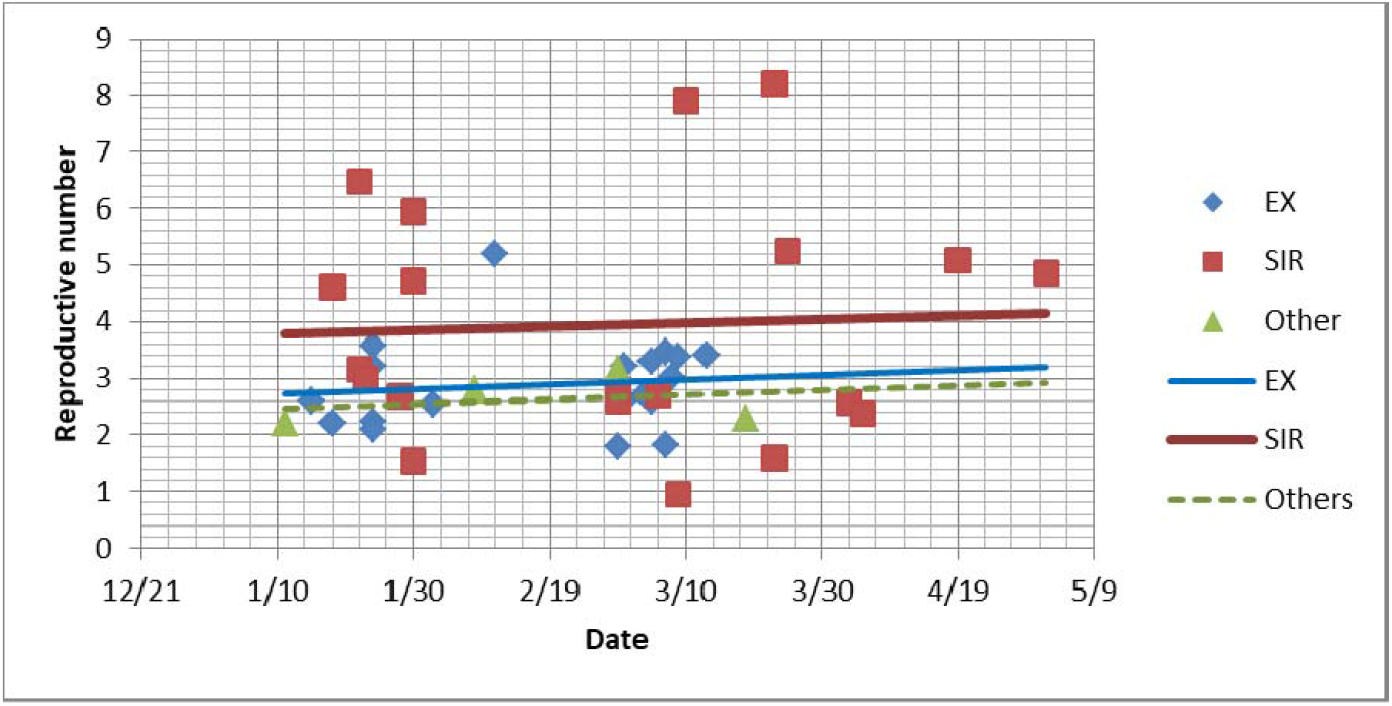
The estimations of BRN based on various methods from the data collected at different time. EX indicates the methods based on exponent-growth-like models and SIR denotes the ones based on SIR models.

The mean and standard deviation of BRN calculated by different classes of method are tabulated in Table 2. It can be observed that the output of SIR methods is higher but more divergent than others. The reason for this is complicated.

**Table 2.** The mean and standard deviation of BRN calculated by different classes of methods.

| Parameters | EX method | SIR method | Others |
| --- | --- | --- | --- |
| Mean | 2.72 | 3.64 | 2.37 |
| Standard deviation | 1.07 | 1.96 | 0.90 |

At the beginning of the break of COVID-19, the data reported by Wuhan in China is less than the ground truth[15, 36, 37]. Note that EX methods are only competent to calculate BRN from early transmission data[8, 38]. In contrast, SIR methods can be applicable to the whole development process of an epidemic and hence more data can be available. For this reason, the EX methods suffer more ill effects of under-reporting than the SIR methods.

On the other hand, for parameter estimation of epidemic models, the SIR methods usually involve nonlinear optimization, which can easily get stuck into improper local minima[39]. Worse, the parameters are frequently estimated at the cost of model simplification. It is hard to obtain a balance between simplicity and practicality. For this reason, the results of SIR methods are relatively unstable. This may provide an explanation of why the output of SIR methods is more divergent than others.

### Transmissibility increases with time?

From Figure 1, it is a consensus that the estimations of BRN increase slightly with time. Does the transmissibility of COVID-19 increase as well? In fact, the possible causes of BRN increase are threefold: (1) behavior changes of population, (2) increase of transmissibility of infectious agent and (3) computational errors. Among them, behavioral changes such as wearing mask typically reduce the probability of transmission and ultimately lower rather than enhance BRN. So, if the third cause is impossible either, it is reasonable that the transmissibility of infectious agent of COVID-19 increases with time.

To explore the reason, we carried out a further investigation into the estimation process of BRN. Among them, we found that up to 27 estimates are derived directly from original data reported by Wuhan authority, which is less than the ground truth due to delays in diagnosis and laboratory confirmation in the early stage [15, 36, 37]. Therefore, under-reporting results in an underestimation, which can give a reasonable explanation for BRN increase. As a consequence, no enough evidence demonstrates the transmissibility increase of infectious agent of COVID-19 throughout the world.

### Magnitude of BRN

According to the aforementioned definition, because *k* and *p* are related to numerous biological, sociobehavioral, and environmental factors in the special geographical location and historical period, BRN *R*_0_ is not a biological constant for a pathogen, or a measure of disease severity[3]. Nevertheless, under the same or similar sociobehavioral and environmental conditions, BRN, as an epidemiologic metric, should be higher for the stronger infectious agents. For this reason, we compared the outbreak of COVID-19 with SARS, as tabulated in Table 3. Obviously, the outbreak size of COVID-19 has already far exceeded SARS. In this sense, the BRN of COVID-19 should be far more than that of SARS, i.e., 2.3-3.6[1, 40].

**Table 3.** The comparison of transmissibility between SARS and COVID-19[41]

| item | Cases in<br>*China* | Control measures in<br>China | Cases in the<br>world | Countries with<br>confirmed cases | year |
| --- | --- | --- | --- | --- | --- |
| SARS | 7,747 | No lockdown | 8,422 | 29 | 2002-2003.6 |
| COVID-19 | 89,526 | Lockdown | 20,423,897 | 212 | 2019-2020.8.13 |

BRN *R*_0_ is originally designed to reflect the characteristic of an infectious agent and hence it is required to be measured from the data in the early stage and in the absence of control measures. In this sense, the data of Wuhan, in which COVID-19 began[42], should be the optimal choice to measure the BRN. On the other hand, BRN is more likely to be closer to the estimation from the data of countries with fewer control measures, such as USA. For this reason, here we focus on the BRN of COVID-19 in USA and Wuhan.

The BRN of COVID-19 in USA is estimated to be 8.21 by Xu et al [6]. In fact, after hearing the dangerousness of COVID-19 from Wuhan, American masses may change their behaviors more or less. Therefore, if COVID-19 spreads from USA, the BRN should be more than the estimation 8.21.

With regard to the BRN of COVID-19 in Wuhan, it is a more complex problem due to under-reporting. Despite a large amount of underestimation appearing in the literature, the several estimations are still similar to the BRN in USA. How to do so? We investigated the data and method, as given in Table 4. It can be observed that most of them utilized compartment models and all of them did not use the raw number of confirmed cases reported by Wuhan authority. Therefore, it is possible to avoid misestimating.

**Table 4.** The data and method of estimation of COVID-19 in Wuhan city.

| Date | $R$ | Data | Model | Predicted end time | First author |
| --- | --- | --- | --- | --- | --- |
| 1/22 | 6.47<sup>#</sup> | Corrected data | SEIR | - | Tang[45] |
| 1/30 | 4.71 | Removed early data | SUQC | late-Mar. | Zhao[16] |
| 2/4 | 2.35* | Confirmed cases in nationals | SEIR | - | Kucharski[46] |
| 2/11 | 5.2 | Confirmed cases in nationals | EX | - | Mizumoto[18] |
| 3/10 | 7.9 | Confirmed cases in nationals | SEIR-HC | 15 Apr. | Zhu[27] |

<sup>#</sup> “control reproduction number”, i.e., reproduction number when control measures are enforced[45].
\* “median daily reproduction number” is the mean number of secondary cases generated by a typical infectious individual on each day in a full susceptible population[46].

Unlike EX methods, SIR methods can be tested by the predicted results. And, Zhao et al[16] and Zhu et al[27] made a prediction about the end time of COVID-19 in Wuhan, as presented in Table 4. The fact is that, on April 15, the last support medical team was evacuated from Wuhan[43] and Wuhan Thunder Mountain Hospital was officially closed on the same day[44]. For this reason, we believe that the model presented by Zhu et al[27] is more reasonable. In addition, the result computed by Tang[45] demonstrates that the BRN in Wuhan should be more than 6.47, since BRN is typically more than control reproduction number. Given these points, there is strong possibility that the BRN of COVID-19 in Wuhan is 7.9.

In summary, the most likely estimation of BRN of COVID-19 in USA and Wuhan is about 8.21 and 7.9. Arguably,it is timely hospitalization that is the reason why the BRN in Wuhan is lower than USA. It is worth noting that limited evidence supports the applicability of BRN outside the region where the value was calculated[47]. For this reason, BRN just reflects the transmissibility of an infection in the special location and period and no BRN is fit for everywhere. To compare with other infections, an alternative term, which is independent of population density, social organization, seasonality and folkway, is desired but still open.

### Findings

The early data is crucial to estimate BRN. Underreporting tends to make an underestimation. In terms of BRN estimation of COVID-19, the SIR-like models are more useful than exponential growth model. The most likely estimation in USA and Wuhan is about 8.21 and 7.9. However, no enough evidence demonstrates the transmissibility increase of infectious agent of COVID-19 throughout the world. Note that it is dangerous to use BRN outside the region where the value was calculated. To compare with other infections, an alternative term, which is independent of population density, social organization, seasonality and folkway, is desired but still open.

## Data Availability

No

## Acknowledgment

This work is supported by National social science foundation of China (16BXW005), Chongqing Basic Science and Frontier Research Project, China (No. cstc2017jcyjAX0007, cstc2017jcyjAX0386), Project Foundation of Chongqing Municipal Education Committee, China (Grant No. 17SKG050).

